# Assessment of benefits of Echo Workflow Solution: Findings from a two-step Delphi study

**DOI:** 10.1101/2022.06.01.22275807

**Authors:** S. Boonyaratavej, F. Riccobene, F. Falahi, J. Franke, J. Solis, R. M. Lang

## Abstract

**Objective:** This study aimed to assess the clinical, operational, and economic benefits of Echo Workflow Solution comprising cardiac ultrasound systems and their advanced echo imaging tools; Ultrasound Workspace tools for advanced visualization and analysis; and the IntelliSpace Cardiovascular platform for image and information management and reporting.

**Method:** A modified Delphi method consisting of two rounds of queries was used to survey a panel of seven experts at geographically dispersed sites (Spain: 2, Thailand: 1, Saudi Arabia: 1, United Kingdom: 2, and USA: 1). Experts were identified via referral and selected based on experience with the solution. Both rounds of the Delphi process were conducted via a verbal or offline questionnaire between December 2021 and April 2022. Participants rated 14 attributes of the solution on a Likert Scale and added qualitative responses.

**Results:** Consensus was unanimous that the solution can enhance access to complete and longitudinal data needed for patient care decision-making; can lead to increased report consistency, measurement reproducibility and comparability; can help with better collaboration within the user’s organization, can help with saving report turnaround time and with improving time-to-detection and/or diagnosis.

The majority of respondents (86%) agreed that use of the Echo Workflow Solution can help increase confidence in diagnosis decision-making; can improve staff experience by providing single screen access to the patient data and lower cognitive load.

**Conclusion:** The study demonstrated that the Echo Workflow Solution can offer clinical and operational benefits. Responses indicated that the solution help with several aspects: reducing time to diagnosis, making it easier to accommodate emergency patients, providing time to perform more advanced studies, and/or increasing time spent with patients. Responses also indicated that the increased consistency and reproducibility of studies can potentially eliminate the need for unnecessary exams, which may result in a better patient experience and economic benefits for healthcare systems. The value of reproducibility, improved time to detection/diagnosis and increased confidence were particularly evident in complex cases and for patients requiring serial studies.

## Introduction

Echocardiography is the most widely used non-invasive imaging technique in clinical cardiology. In fact, its availability, reliability and cost-effectiveness, combined with its lack of ionizing radiation and non-invasive nature, often render it the first-choice imaging technique for the diagnosis and follow-up of most heart diseases.^1^

Increased use of echocardiography has been accompanied by the development of systems that store tremendous amounts of echocardiography data. While this data is valuable in informing diagnosis and treatment and providing a more complete patient record, it can be highly resource-intensive and time-consuming to access. As data accumulates, connectivity and interoperability among systems become increasingly important so that data can be accessed wherever it is needed, and there is a need for solutions that structure data so that it can be used effectively in research and clinical decision-making.^2^

In addition, the increased use of echocardiography places a high workload on cardiologists and sonographers.^3^ This is driving the demand for more efficient workflow and for improving the staff experience, which is one of the key components of the Quadruple Aim (QAim).^4^ QAim is a framework developed to optimize healthcare system performance. It adds improving the staff experience to the initial goals of improving clinical outcomes, enhancing patient experience, and reducing costs. (Figure 1).

**Figure 1.**
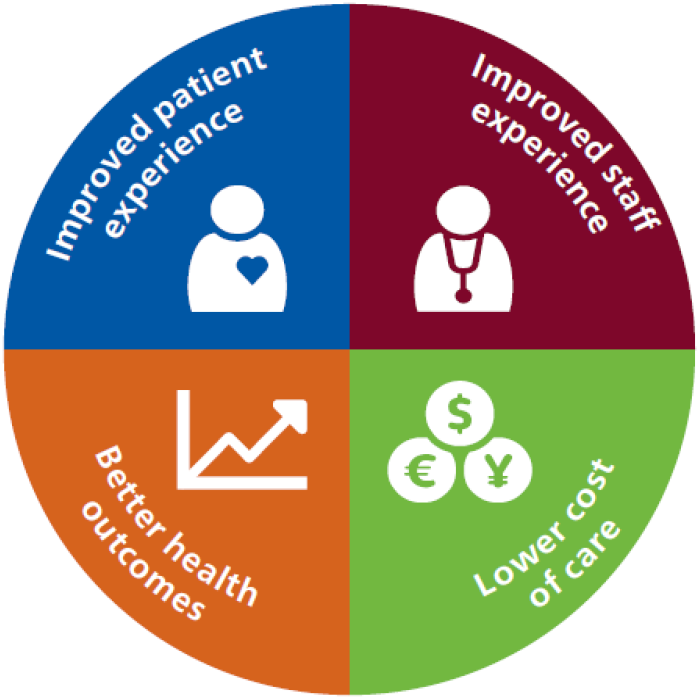
The Quadruple Aim.^4^

All these trends have provided the impetus for solutions focused not only on improving image quality, but also on data access, data integration, interoperability, and workflow. From image acquisition through postprocessing and data retrieval, opportunities exist to improve workflow efficiency and present data in ways that support patients throughout their healthcare journeys, including accurate and timely diagnosis, follow-up and patient management.

Understanding how and to what extent available technologies can help address echocardiography challenges will help manufacturers refine existing products and develop new ones (if needed) and will enable healthcare providers to choose technology that will enhance echocardiography department operations. Therefore, this study was developed to assess the benefits of the Echo Workflow Solution (Philips, the Netherlands) by acquiring information on its real-world use.

The Echo Workflow Solution includes three products that combine to form an end-to-end echocardiography solution:

1. Cardiac Ultrasound (EPIC CVx and Affiniti CVx; Philips), used to acquire images of the heart and perform measurements
2. Ultrasound Workspace (TTA2.5X; Philips), used to provide automated and reproducible image measurements
3. IntelliSpace Cardiovascular Information System (ISCV 2.x/ 5.x/ 6.x; Philips), used to display previous and current cardiovascular images (echocardiography, computerized tomography (CT) and magnetic resonance imaging (MRI)) and patient data through a single point of access, and to provide advanced clinical tools for comprehensive analysis and reporting

## Methods

We used the Delphi method in this study. The Delphi method is a structured process to develop consensus among a panel of experts.^5^ Developed by RAND in the 1950s, it has been used in many industries. Delphi panels may consist of 3 to 80 members who are knowledgeable about the field of study and commit to multiple rounds of questions.^6^ Several rounds of consultation allow for clarification and the correction of potential bias in the initial responses which is the main fragility of classical queries. The Delphi method requires that participants are queried individually, rather than in a group and their individual responses remain anonymous. These requirements encourage critique and prevent the personality or reputation of a participant influencing others.

In this study, a multi-site Delphi method consisting of two rounds of queries was conducted. Both rounds of the Delphi process were conducted via video conferences and paper questionnaire between December 2021 and April 2022. The first round followed a standardized questionnaire in which participants scored 14 attributes of the solution on a Likert Scale, rating their level of agreement with statements on a 1-5 scale. Participants also provided qualitative responses that explained their ratings and added context. The second round sought clarification and elaboration of first round answers.

### Participant recruitment and demographics

Experts in geographically dispersed sites (Spain: 2, Thailand: 1, Saudi Arabia: 1, United Kingdom: 2, and USA: 1) were identified via referral from Philips clinical specialists and selected based on their expertise in echocardiography and their experience with the Echo Workflow Solution and its components. All participants are cardiologists and/or head of echocardiography lab who are daily users of the solution with 7 to 30 years of experience in echocardiography. (Table 1)

**Table 1.** Experts demographics (n=7)

| Characteristic | Number of Experts |
| --- | --- |
| Years of experience |  |
| <10 | 2 |
| 10-19 | 3 |
| >20 | 2 |
| How often do you use the solution? |  |
| Daily | 7 |
| Other | 0 |
| Affiliations |  |
| Affiliations with an academic center | 6 |
| President of national/international cardiology societies | 1 |
| Member of cardiology societies | 6 |

### Data analysis

Quantitative data for questions was analyzed using Microsoft Excel 365. Analysis of round one questions demonstrated the need for clarification on some items, which was obtained in round two. Round two interviews also focused on deeper exploration of the clinical impact of the solution. All data was collected prospectively and analyzed using Microsoft Excel to obtain descriptive and summary statistics.

### Ethical considerations

The results presented in this article were derived using the modified Delphi Panel approach, which is based on expert opinion. All participants signed informed consent agreements and no individual can be identified from the results. This study did not require any patient data, so no patient data was collected, and no patient samples or tests were used.

## Results

The study demonstrated that the Echo Workflow Solution can help with clinical, efficiency and workflow benefits as described in this section. Note that in the summaries of results provided below, the term “agreement” refers to answers of both “completely agree” and “somewhat agree,” and “disagreement” refers to “completely disagree” and “somewhat disagree.” Anonymized responses to each question from each respondent are available as supplementary data (Figure 4 and supplementary data).

### Clinical benefits

Four statements addressed clinical benefits, including decision-making confidence, time to diagnosis, consistency and reproducibility, and clinical collaboration (Figure 4 and supplementary data).

#### Decision-making confidence

Full consensus was achieved that use of the Echo Workflow Solution has led to increased confidence in diagnosis/decision-making. The experts who expanded their responses noted that the solution helped with increased confidence by offering exam quality, access to information, reproducibility, the possibility of reviewing previous exams, and the ability to create and access comprehensive and structured reports.

In the second round, clarification was sought concerning if improved confidence in decision-making is more significant for specific clinical indications. One stated that, “*Decision-making has been enhanced [using the solution] in any scenario which needs several measurements, and/or requires consistency and comparison with previous measurements. Examples are valvular heart disease, adult congenital disease, and cancer patients*.” One noted, “*You must be a good cardiologist [to make the accurate diagnosis decision], but you also require good tools to support you. This solution helps me to be confident about complex cases*.” Another stated that, “*The more complex the scenario, the greater the benefit of this solution*.”

#### Time to detection/diagnosis

The second clinical benefit statement, concerning if using the Echo Workflow Solution had led to improvements in time to detection and/or diagnosis, also achieved 100% agreement. Those who expanded their responses noted that access to all necessary tools that help with following the clinical guidelines, higher quality images and advanced analysis may mean fewer tests are required to reach a definitive diagnosis and therefore diagnosis may be faster.

The second-round questions further explored the cases for which improving time to diagnosis makes a significant impact (Figure 2). An expert highlighted that, “*Time is critical for fast diagnosis for intensive care patients and complex patients (for example, valvular heart disease patients when there is more than one valve involved and patients with pulmonary hypertension, as sometimes the right ventricle is very complex, and we need to use all the tools in our system). Cancer patients are not complex cases but sometimes you have the echo in the morning and the patient sees the oncologist on the same day, so the report should become available quickly*.” In addition, one mentioned the solution can help with consultation and second opinions. For example, it helped them provide consultation during the pandemic when pneumonia or pulmonary edema was suspected in COVID-19 patients.

**Figure 2.**
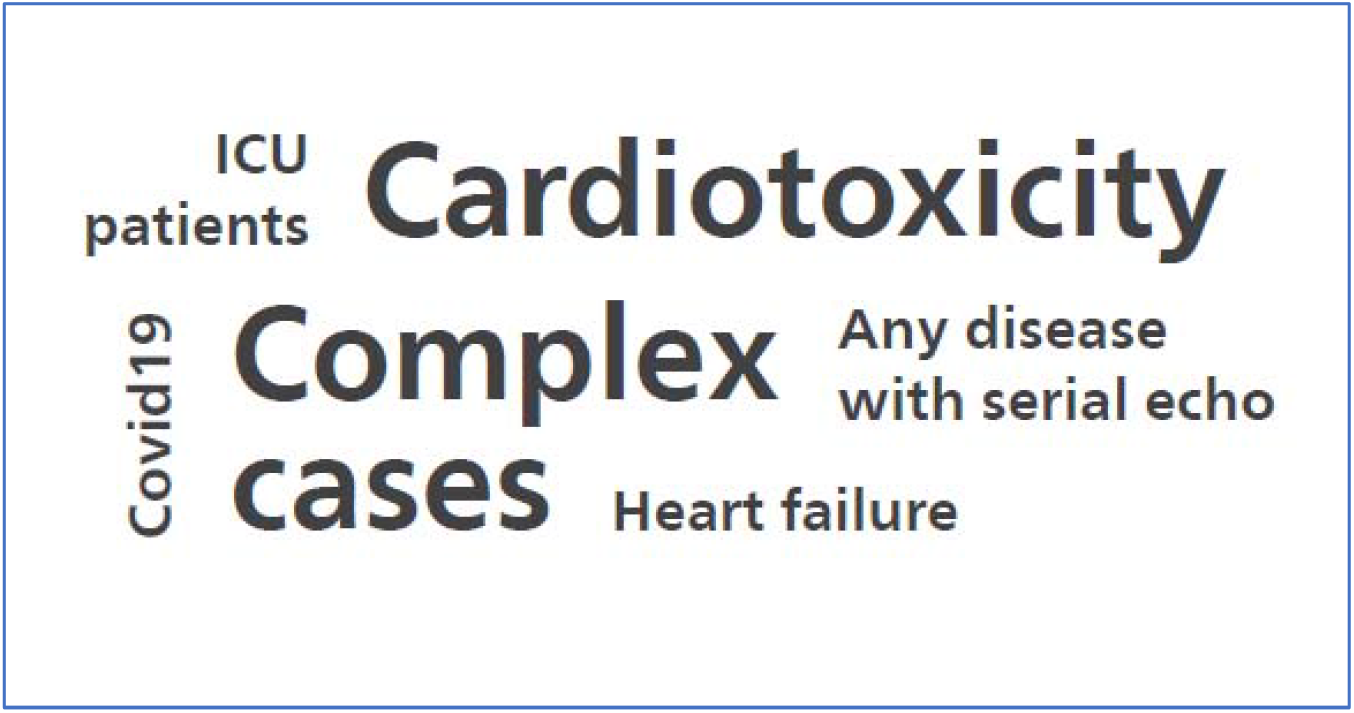
Clinical indications named by survey participants as particularly benefiting from improved time to diagnosis/detection, as one of the benefits of the Echo Workflow Solution.

#### Increased consistency and/ or reproducibility

All seven respondents completely agreed that the Echo Workflow Solution has led to increased report/measurement consistency &/or reproducibility. Two respondents noted the value of comparison with the previous data when interpreting new data, and one stated that a critical aspect in reporting is consistency.

Respondents mentioned several clinical scenarios benefiting from the increased consistency/reproducibility offered by the solution (Figure 3). They noted that reproducibility is particularly crucial for patients who require serial measurements. This includes patients with cardiotoxicity, because any change in values can necessitate a change to their cancer or cardiac treatment, and valvular disease patients, who may require several follow-up exams.

**Figure 3.**
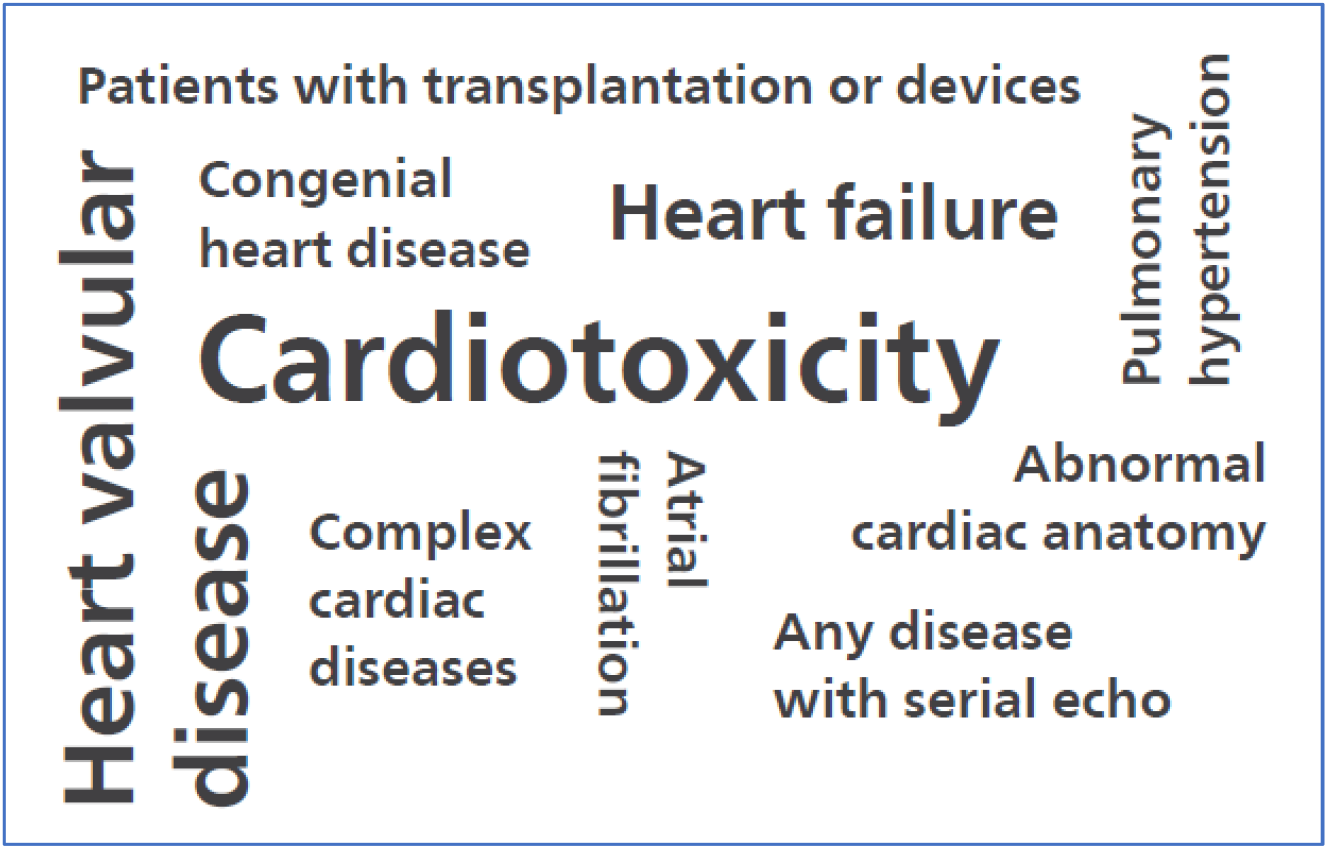
Clinical indications named by survey participants as particularly benefiting from measurements/report consistency and comparability, as one of the benefits of the Echo Workflow Solution.

#### Clinical collaboration

All participants agreed that the Echo Workflow Solution has led to better collaboration within their organization. Respondents noted that the solution helps discussion during multi-disciplinary meetings, makes it easy to review and share the report within the organization and enables easy transfer of images and reports to other departments.

### Patient experience

When judging if the Echo Workflow Solution can improve the patient experience, 86% agreed, and 14% disagreed (Figure 4 and supplementary data). A respondent who disagreed noted that “*It’s a general issue in [my] country, because the physician who orders the echo does not have access (immediately or easily) to the patient data in [our] electronic health record system to view if an echo was done [and to avoid an unnecessary exam for patient]*.” Among the potential benefits to patients that were noted were a decrease in repeat exams because of access to previous exams, shorter exams that can allow patients more time with the doctor, and patient confidence that the institute has all the patient’s images. Two respondents shared opinions about which patients benefit most from the solution. One responded, “*Inpatients who are admitted in the hospital and need repeated exams in a short period of time benefit. It is very important to have all images and to be able to compare them and avoid unnecessary repeated exams*.” The other noted, “*One ICU patient had severe pulmonary arterial hypertension and the patient’s oxygen saturation dropped. One possible explanation for this is PFO (Patent Foramen Ovale). I reviewed a previous exam that clearly showed PFO, so there wasn’t any need to repeat the examination. This led to faster diagnosis*.*”*

**Figure 4:**
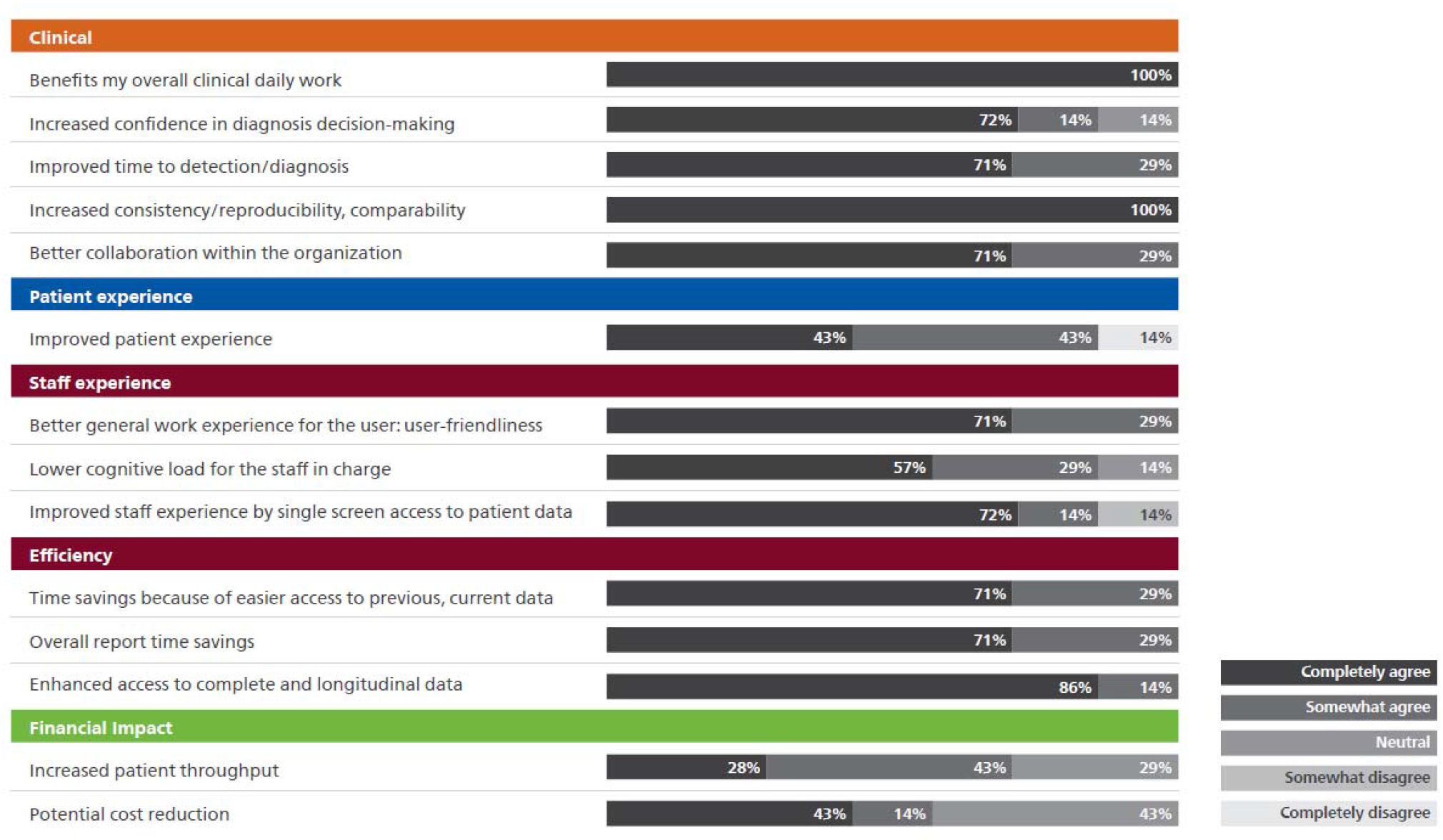
Responses to 5-point Likert-style questions. Participants were asked to assess the Echo Workflow Solution in comparison to a situation in which they did not have the full Echo Workflow Solution. Percentages are rounded up to the nearest digit.

### Staff experience

The questionnaire included statements that addressed staff experience in three areas: general work experience, cognitive load, and single-screen access (Figure 4 and supplementary data).

Consensus was reached that the Echo Workflow Solution has led to better general work experience for the user. Qualitative answers included that the solution helped their overall experience, was user-friendly immediately or after a few weeks of use, and that the user-friendliness of the system helps residents learn new tools quickly. When asked if the Echo Workflow Solution has led to lower cognitive load for the staff in charge, 86% agreed and 14% were neutral. In response to a statement about the Echo Workflow Solution leading to improved staff experience by single screen access to the patient data,” 86% agreed and 14% disagreed. Participants noted that there was now a single environment for all data, there was “continuity,” IntelliSpace Cardiovascular played an important role as it provides access to cardiac CT and MRI images in addition to echocardiography, and that EMR (electronic medical record) data and imaging data is on the same screen. Two participants stated that at their facilities, the EMR data was not yet integrated or not fully integrated.

### Efficiency: Time savings

Time savings was assessed in terms of both report turn-around time and savings due to easy access to data (Figure 4 and supplementary data).

We achieved consensus on the statement about the Echo Workflow Solution leading to overall report turnaround time-saving. All experts agreed they experienced time-saving by having easier access to previous/current multiple images and results. One expert stated that because an exam can be viewed even if performed at a different location, there is less need to repeat exams and it saves time. Another noted that they see an immediate time-saving and this savings allows more time for reviewing images, postprocessing, and to use tools that they wouldn’t have time for otherwise.

### Healthcare financial impacts

Participants were queried about two potential healthcare economic benefits: patient throughput and organizational cost reduction (Figure 4 and supplementary data).

This study sought to uncover if the Echo Workflow Solution affected the number of patients that could receive exams in a day. Of those surveyed, 71% agreed and 29% were neutral or said the statement was not applicable to their hospital setting. Some noted that the solution provided flexibility to accommodate urgent patients and do higher quality studies. One stated, “it *[the solution] helps with workflow plasticity: being flexible and adaptab*le.”

Concerning if the solution and its benefits, such as efficiency improvement and diagnosis accuracy, might help the participants’ department/institution to lower costs, 57% agreed and 43% were neutral. One noted that saving time is an indirect cost saving, because “*time is money*.” Among the positive comments were, “*Time saving is one aspect; however, one important aspect is the outcome of the patients. A solution or system that allows you do a better, faster diagnosis does help patients eventually. That is cost-saving, as patients are diagnosed quicker and get treated quicker, so helps with long-term cost saving*.” *“It makes staff have a better experience and therefore it might help with staff retention*.” “*Potentially we can use data to reduce the number of (unnecessary) echoes performed; many are unnecessary*.*” “It [the solution] is worth the investment, [and is] necessary for the echo lab*.”

## Discussion

Areas of consensus suggest that the Echo Workflow Solution can contribute to clinical benefits, workflow efficiency and a positive staff experience, three areas that are essential to Echo lab.

Regarding clinical benefits, this study’s qualitative responses implied that high-quality exams and reports and efficient access to patients’ data as offered by the solution can help with improving time-to-diagnosis and diagnosis decision-making. The cardiologists participating in this study highlighted that the improving time-to-diagnosis and diagnosis decision-making offered by Echo Workflow Solution can be more significant for the complex cases or for scenarios that require several measurements over time. For instance, measurement consistency and reproducibility and access to longitudinal data were deemed especially valuable when they provided a basis for comparison in treatment assessment. Access to longitudinal data may also reduce unnecessary repeat exams. This is significant because unnecessary exams represent a cost burden not only to the healthcare system, but also to the patients in some countries. One study of 34,600 patients who received imaging found that close to 7.7% of the patients had additional images taken for the same condition within 3 months due to lack of access to previous images.^7^ Decreasing unnecessary repeat exams can also potentially improve the patient experience, because patients are not subjected to exams that do not add clinical value. Although Echo Workflow Solution may support to some extent, cross organizational data integration is crucial to fully achieve the goal of reducing unnecessary exams.

In addition to clinical benefits, cardiologists need systems that also help efficiently manage echocardiography workflow. This study suggested that the Echo Workflow Solution can help with saving time both in accessing data and in reporting. When workflow is faster, it may provide time to perform more comprehensive echo exams. As one respondent explained, “*For a certain type of patient we can do studies with more quality (more 3D echo, more strain) that could not be performed previously because of the lack of time and absence of necessary tools. Some studies could not be done previously, because additional analysis which takes a few more minutes could not be done as frequently*.” Although 100% of the respondents agreed that the solution helps speed workflow, fewer (71%) agreed that time savings leads to increased patient throughput, because patient throughput relies on several factors, including the number of sonographers, echo machines and rooms, which are outside the influence of the Echo Workflow Solution. Yet having a fast and reliable solution allows staff to accommodate emergency patients and perform more advanced analysis (supplementary data).

Finally, participants’ responses revealed that the Echo Workflow Solution can help improve the staff experience. This can be a result of, for example, less manual work to enter data into the report, less time spent looking for data, workflow flexibility, and having efficient access to data. This study’s findings complement studies that have shown that data integration, intuitive interfaces, reduced documentation, and fewer manual tasks help reduce user fatigue and decrease burnout.^8,9^

This study was conducted to understand to what extent the Echo Workflow Solution can help the Echo lab in the context of an integrated workflow and what needs to be further improved therefore the findings may be of interest to cardiologists specialized in echocardiography, sonographers, Echo lab management, and researchers. It is important to note that participants of this study in multiple geographical locations practicing in different healthcare systems and with varied patient populations found value in the Echo Workflow Solution. This is a particular strength of the study because it indicates that the solution can be beneficial in various healthcare systems. While the Delphi method is an excellent method for measuring consensus, the next step is to quantify and measure some of key benefits over time.

## Data Availability

All data produced in the present work are available as supplementary file

## Data availability

All questionnaires with all anonymized answers given by Delphi panel participants are available as supplementary data. This data was used to generate figures and tables in this article.

## Acknowledgement

We would like to thank the following members of the Philips team whose efforts facilitated the success of this document.

1. Sergio Garcia Casado; Cardiology Informatics Senior Consultant
2. Chantra Sukyon, Business Manager EDI, Philips Thailand
3. Hui Li Kim; Senior Business Marketing Manager, Cardiology Informatics, Philips APAC
4. Parastou Islami; Clinical Scientist
5. De Wet Nel; Ultrasound Clinical Applications Specialist

