## Supplementary data for "Assessment of benefits of Echo Workflow Solution: Findings from a two-step Delphi study"

*“The aim of this survey is to know the benefit perception of the echo workflow for your institution from different angles: diagnosis benefits, efficiency benefits and workflow improvements. The echo workflow solution is created from the combination of different parts which creates a single cardiac echo pathways workflow.*

*The parts to take into account within the echo workflow are: Echo Carts and its advanced echo imaging tools, the Intellispace Cardiovascular platform and its workflow, reporting and viewing capabilities and the TomTec tools for postprocessing advanced imaging which is part of this platform.*

*All questions are related to the combination of those three parts and the integrated workflow that derives from it.”*

Question 1. Please describe your function and role

| **Comments** | |
| --- | --- |
| 1 | Cardiologist in Echocardiography lab; Echocardiographer; 15 years experience  My role is to acquire and report the daily work in echocardiography, mostly transthoracic, and transesophageal Echocardiography; Also Interventional echocardiography monitoring, surgical echocardiography support. Also in other areas needing my input CCU, Emergency in (mainly cardiovascular area but also many other departments); Co-lead the echo lab: coordinator of the research in echocardiography lab, one of the top experts in Echolab |
| 2 | Chief Echo cardiographer |
| 3 | Director Noninvasive Cardiac Imaging Laboratories; Heart & Vascular Center |
| 4 | Head of Echo department; management and clinical participation; responsible for training, service delivery, etc. Experience: 8 yrs; Member of national Society of Echocardiography |
| 5 | Director of adult Non-invasive cardiovascular lab; Head of division of cardio-vascular medicine; President of [national] Society of Echocardiography |
| 6 | Director of noninvasive imaging department with the focus on Valvular Heart Disease. At our hospital, the echo lab is used for complex Echo (e.g. for aortic stenosis) |
| 7 | Registrar of Cardiovascular Medicine; 11 years of experience in echocardiography |

Question 2. How often are you using the Echo workflow solution?

| **Response** | **Number** | **Percent** |
| --- | --- | --- |
| Never | 0 | 0% |
| **Daily X** | 7 | 100% |
| Monthly | 0 | 0% |
| Not Sure | 0 | 0% |
| Other | 0 | 0% |

Question 3. Please describe the type of patients you most frequently use the Echo workflow solution with (including information on the age, sex, admission diagnosis, comorbidities, and medications).

| **Comments** | |
| --- | --- |
| 1 | General cases: Cardiovascular patients- all the patients with coronary heart disease or heart valvular disease- typically men and women and usually patients above 60yrs old;  Specific cases: We also have a large congenital heart disease unit (ref unit in the country) in this center there is a specific subset of who are younger patients with complex heart diseases, these are patients needing a lot of echocardiography test during their life-time.  -Cardiotoxicity patients in our hospital are: mainly breast cancer in our hospital (typically younger than cardiovascular patients) and hematology disease patients: leukemia and lymphoma patients who require high number of echocardiography more echo test and serial testing  Can a fast workflow be beneficial for the cardiotoxicity patients than other patients? Can this subset of patients clinically benefit from a fast echo workflow?  Yes. Cardiotoxicity patients usually require a very focused exam, in which the same feature (left ventricular systolic function) needs to be assessed very accurately, whereas other aspects are mostly normal or with minor abnormalities. For this reason, focused acquisitions, postprocessing (strain) and reporting using a fast workflow may be the best solution. |
| 2 | A wide range of patients; Adults >18 we can do above 16; Gender varies mainly male 60-40  LV Ejection fraction, post myocardial infarction. We have cancer patients (Cancer patients are probably 20 %) where we use the most GLS and analysis for ejection fraction, we use either Ultrasound Workspace or dynamic heart model. we are considered a big cardiac thoracic centre. The majority are cardiology patients for e.g. preop-, post op assessment of the valves, post MI. we have heart failure team where we include the cancer patients. Comorbidities: hypertension, diabetes, hyperlipidaemia |
| 3 | We cover adult patients > 18 with any diagnosis, any gender, medications etc. 120 Echos a day or so. Screening, diagnosis, very complex patients. It’s a wide range of indication.  We have a large transplant program one of the biggest in the country so a lot of Echos in transplant, but overall, we cover any type of indication. |
| 4 | A wide range of patients; Adults; e.g. Cancer patients and more female 40-60 |
| 5 | A wide range of patients; Age: Adult patients >18; wide range also patients 100  Gender: 50:50; Young patients: congenital or rheumatic heart disease; Adult patients: coronary artery disease or degenerative cardiac disease e.g. Aortic Stenosis, Transvalvular aortic valve replacement (TAVR); Structural interventions: Pre and Post procedure  Complex cases: we are referral center and receiving patients with different comorbidities from other centers |
| 6 | Adults mean age is 40-100; The mean is 40 (Pediatric, also 70, also cancer patients); Complexity is high at our hospital (maybe the most complex hospital in my country): many hospitals refer patients with valvular heart disease, Heart failure, Pulmonary hypertension; Cardio-oncology; |
| 7 | Age: adults more than 18 years old  - Gender: both male and females  - Clinical scenarios: all cardiac complaints, and diagnoses |

Question 4. Does the Echo workflow solution improve or support your work? If YES, how

| **Response** | | **Number** | **Percent** |
| --- | --- | --- | --- |
| Access to data | | 7 | 100% |
| Sharing of results | | 6 | 86% |
| Time-saving in report preparation | | 7 | 100% |
| Consistency of the report and reproducibility of exam measures | | 7 | 100% |
| Quality of report | | 7 | 100% |
| **Comments** | | | |
| 1 | Certainly, accessing to previous data of patients  Many of these patients the cardiovascular but also the subset of patients need repeated testing -> some of them require a lot of information in the report, the workflow certainly saves a lot of time  high consistency for the report- as you can use the previous report and only adding additional information, so the report gets more and more complete every time; structural reporting helps with higher consistency)  you can measure the quality of report based on whether the report includes the minimum data needed for decision making in patient, quality of the report withing this workflow solution has improved | | |
| 2 | The top one is the consistency- e.g. for cancer patients you need highly accurate measurements and you should be able to with previous exams to monitor the function of heart- therefore quality of images and consistency of the report as well to monitor patients. | | |
| 3 | When patient comes, I can access the old study and compare it. | | |
| 4 | - | | |
| 5 | Sharing of results (important for communication with colleagues; sharing results with interventionist and CVT (Cardiovascular Thoracic Surgeons)  We have patients with chemotherapy; oncology department ask for Left ventricle (systolic function); we perform the echo, and the oncology department can look at the results This helps to streamline the workflow for a patient not only for cardiologists but whoever takes care of patients.  Time-saving in report preparation (time is well spent) when you can send over the structured report from echo cart; all the numbers into ISCV and you can do the interpretation and making report. Quality of report its easier to do quality control by having all the information in one system and you can look at it systematically. Uniformity of the report is also the benefit | | |
| 6 | time saving is perhaps the most important because he can spend more time with the patients | | |
| 7 | - | | |

Question 5. Please state to what extent you agree with the following statement: The Echo workflow solution has led to **enhanced access to complete and longitudinal data** needed for decision-making (compared to not having the Echo workflow solution)

| **Response** | | **Number** | **Percent** |
| --- | --- | --- | --- |
| Completely disagree | | 0 | 0% |
| Somewhat disagree | | 0 | 0% |
| Neutral | | 0 | 0% |
| Somewhat agree | | 1 | 14% |
| Completely agree | | 6 | 86% |
| **Comments** | | | |
| 1 | Currently cardiology department has access to this type of data, but many of the physicians receiving our report are outside of cardiology department-> not all the final receivers of report have access to the longitudinally data. | | |
| 2 | - | | |
| 3 | - | | |
| 4 | - | | |
| 5 | - | | |
| 6 | - | | |
| 7 | - | | |

Question 6. Please state to what extent you agree with the following statement: The Echo workflow solution has led to **increased report reproducibility/consistency, comparability, (and observer independence)** (compared to not having the Echo workflow solution)

| **Response** | | **Number** | **Percent** |
| --- | --- | --- | --- |
| Completely disagree | | 0 | 0% |
| Somewhat disagree | | 0 | 0% |
| Neutral | | 0 | 0% |
| Somewhat agree | | 0 | 0% |
| Completely agree | | 7 | 100% |
| **Comments** | | | |
| 1 | The report reproducibility has certainly enhanced because the solution gives you the tools that you need every time (comparable); all users in the unit work with the same framework a junior physician is able to review previous studies from a senior specialist; the technology allows you to look at previous data in order to interpret your own data -> homogeneity in the interpretation  I am not sure about the observer independence, but Comparability between studies which is important has improved.  Can these benefits (comparability of studies and increased report reproducibility) be considered as clinical benefits?  Yes. A critical aspect in reporting is **consistency**. When the clinician has to interpret two serial studies from the same patient, comparability and consistency is key. | | |
| 2 | It’s a benefit for everyone regardless of their experience in the field | | |
| 3 | Reproducibility helps a lot. The way we work today forces every physician to write the report similar. | | |
| 4 | Reproducibility has a direct clinical impact. Sonographer and interventionist benefit from it- you can see previous data which is key. | | |
| 5 | Personally, I completely agree with increased report reproducibility and comparability.  Technology may play a role with reproducibility – example: when the cardio oncology patients who have chemotherapy come for follow-up echo, it would be very helpful (during acquisition) to pull up the previous exam to compare the exams side by side. | | |
| 6 | Clinicians working with us are happy with their report because variability decreased with this solution. Because we have tools that we trust and even if it is manual, it is reproducible.  Automatic tools such as Autostrain increase the reproducibility. We are confident in the tools and trust the results. We use the solution for diverse patients e.g. in valvular hearts disease and heart failure as mentioned in the guidelines. | | |
| 7 | - | | |

Question 7. Please state to what extent you agree with the following statement: I have experienced **overall report turnaround time-saving** in my work due to the use of Echo-workflow solution (compared to not having Echo workflow solution.)

| **Response** | | **Number** | **Percent** |
| --- | --- | --- | --- |
| Completely disagree | | 0 | 0% |
| Somewhat disagree | | 0 | 0% |
| Neutral | | 0 | 0% |
| Somewhat agree | | 2 | 29% |
| Completely agree | | 5 | 71% |
| **Comments** | | | |
| 1 | Certainly TAT decreased- saving a lot of time in different steps of the workflow is clear- e.g. we don’t work much on the machine and we used to take a lot of measurements and do a lot on the machine, now it has been decreased; speeding up the work -> patients can be out quicker and new patients can come quickly  Reporting time, Image analysis more quickly than before | | |
| 2 | We have 45 min slot to see the patient we do comprehensive study, with 3D. lots of data is acquired; we spend 20 min scanning the patient and 20-25 min reporting. Probably in the past without the solution: the Echos were not as much comprehensive and not such amount of data could be acquired. now with advanced Echos so many things can be done. I can say time efficiency is better now. If we did not have these tools, it could have taken us very long time to report. | | |
| 3 |  | | |
| 4 | ISCV prefilled that otherwise you must write manually. | | |
| 5 | - | | |
| 6 | The time from acquisition to reporting: By having 3D and different tools, automation (Autostrain), structured report we could save time (reduce scan time about 50%). Measurements go to the report which save time too. The workflow is easy using this solution. | | |
| 7 | - | | |

Question 8. Please state to what extent you agree with the following statement: I have experienced **time-saving because of the easier access** to any previous, current/ multiple images &/ results (compared to not having Echo workflow solution).

| **Response** | | **Number** | **Percent** |
| --- | --- | --- | --- |
| Completely disagree | | 0 | 0% |
| Somewhat disagree | | 0 | 0% |
| Neutral | | 0 | 0% |
| Somewhat agree | | 2 | 29% |
| Completely agree | | 5 | 71% |
| **Comments** | | | |
| 1 | Previously exams were saved individually, access to previous studies in the past was a challenge.  ISCV provides immediate access to previous reports of the patient´s records. However, access to images depends on the institutional PACS communication with ISCV, that in some instances slows down the workflow. This is more a technical issue with the institutional PACS storage management. | | |
| 2 | You can have multiple images and you can look at previous exams these helps to save time. | | |
| 3 | Happy with the speed of finding older studies. | | |
| 4 | If you don’t have the report this way, you must try to find it from somewhere.  It's an immediate time saving. Once you get more time, you can do more measurements, postprocessing, and you can use the tools that you would not use if you are under so much time pressure.  By a faster workflow, you can take more time on postprocessing and looking at the images a bit more or better which would reflect on the patient. | | |
| 5 | By having easy access, you can view images from different location than the exam was done; e.g. patients admitted to CCU, it helps to view the previous images if the previous one has good quality and sufficient information, no need to repeat the echo.  In my institution we look at the entire system cardiovascular solution not only for the Echo. It helps with echo but also beyond the echo. | | |
| 6 | In radiology and cardiology, it is key to have access to previous data and compare. The more complex is pathology, the more important is to compare. Patient is complex you have to always check the previous data. By saving time we have more time to compare, and accessibility is key for such comparison.Comparing helps with better decision for diagnosis. | | |
| 7 | - | | |

Question 9. Please state to what extent you agree with the following statement: The Echo workflow solution has led to **Improvements in time to detection and/ or diagnosis** (compared to not having Echo workflow solution):

**Clarification note: In this question we refer to Time to diagnosis and not improving the diagnosis itself. In this question we do not refer to better diagnosis itself- The diagnosis itself depends on expertise of the clinician and other factors**

| **Response** | | **Number** | **Percent** |
| --- | --- | --- | --- |
| Completely disagree | | 0 | 0% |
| Somewhat disagree | | 0 | 0% |
| Neutral | | 0 | 0% |
| Somewhat agree | | 2 | 29% |
| Completely agree | | 5 | 71% |
| **Comments** | | | |
| 1 | Now that the workflow is faster, we are doing more echos, we are decreasing the time to detection/diagnosis  Diagnosis of the patient is not exclusively about what the patient has now but also to determine if the condition of the patient is stable or has changed- so the availability of previous studies answers the question if the condition has changed or not.  Also, If you can perform more echos, it can help to exam more patients who are waiting for an exam => the sooner the exam is done, the faster diagnosis will be so it will be shortening time to diagnosis | | |
| 2 | For cancer patients, we can detect earlier using new tools (like GLS)  Avoiding unnecessary tests, avoiding a patient doing an unnecessary MRI | | |
| 3 | Having previous images help. | | |
| 4 | For the patients who are waiting, potentially, yes. However, it means shorten the slot time to accommodate more patients which is not always possible.  We are a big centre with a lot of pressure to accommodate more Echos on a day, I might be able to manage to accommodate an urgent patient in my systems because I have a solution allowing me to be faster with e.g. reporting.  Potentially it can help with improving diagnosis too. (it does not change diagnosis itself but getting to the diagnosis will be faster). Because if you get higher quality images or having advanced analysis, sometimes another test is not required to do the diagnosis. | | |
| 5 | Previously without Echo workflow solution: patient comes in and you have to look at the echo ourselves and allocate the time for coming to the lab personally; it may delay the time of appointment. But with this solution, you can send the patient to the lab and ask a sonographer or a cardiologist to do the echo and you can open it in a different location; this can save time to diagnosis.  If my fellow has question about the echo or not sure about diagnosis or what to do next; consultation is much easier-> they can open the ISCV from anywhere in hospital (and hopefully in future from home is not what we are doing currently)  Consultation remotely is possible. My hospital is one of the few you can open the echo in HIS and that helps because it can easily be used during discussion and teaching. | | |
| 6 | Having various tools makes it easier to get the diagnosis. You can use the tools relevant for indication and type of problem of your patient. To make a decision for heart failure, you refer to the guidelines, where for each step you need a tool, when you have all tools, you can make a faster and better decision. Having new tools to analyze the 3D and morphology lead to faster diagnosis of mitral valve disease as an example.  These tools are good for any doctor making the decision regardless of the years of their experience. For residents, these tools are good for teaching. | | |
| 7 | - | | |

Question 10. Please state to what extent you agree with the following statement: The Echo workflow solution has led to **increased confidence in diagnosis decision-making** (compared to not having the Echo workflow solution)

| **Response** | | **Number** | **Percent** |
| --- | --- | --- | --- |
| Completely disagree | | 0 | 0% |
| Somewhat disagree | | 0 | 0% |
| Neutral | | 1 | 14% |
| Somewhat agree | | 1 | 14% |
| Completely agree | | 5 | 71% |
| **Comments** | | | |
| 1 | Decision making is very important, because the decision making is not exclusively about the patient having a condition.  Over (an estimate) 50-60% patients going to the lab have a previous exam  an important Question for decision making is whether the condition of the patient from previous study has changed or not.  In the past there was a report in the EHR or some images somewhere, now it is very straightforward you can more quickly go to the decision making because now all info in front you -> decision making is  In our experience, structured reporting has contributed to a more comprehensive report in terms of number of parameters and consistency of parameters being reported. All these data provide a stronger support for interpretation. | | |
| 2 | It benefits everyone. The younger staff with much less experience may benefit more in their diagnosis decision. | | |
| 3 | When we are looking at previous studies we make comments, e.g. strain is worsening having all information readily available helps with the decision making. However, doctors who read the report don’t have the time to look at all previous reports. They get only 15 min to finish the work. | | |
| 4 | It benefits everyone.  The solution helps juniors more, because its user friendly and having the data available, easy to share the case with someone to review if they are not in the same place with you  However, the more advanced imaging does help the seniors more because juniors have less experience in interpreting the advanced imaging. | | |
| 5 | This solution helps with reviewing the exam; by reviewing the exam you may pick up something that you missed in the first place.  We have to decide what to do next in the patient journey (surgery, intervention?) Usually in complex cases, you do the conference and CVT, cardiologist looking at the same data during the conference and make the best decision for the patient.  Conferences happen weekly (then we have some additional meetings for emergencies patients) Like internal medicine, they do weekly meeting outside of cardiology discussing patient with e.g. infective endocarditis, in the meeting they just pull up the web-based tool and look at the echo, we can explain to them the meaning and assess the risk, we can look at hemodynamic.  We have various meetings, we have the morbidity mortality meeting, we are looking at the echo and assess the outcome of surgery. Image and data are there to make the best conclusion.  Growing together and helping each other -> make the decision for treatment of patients | | |
| 6 | Because of quality, access to information, reproducibility; All the information is available for the team to make the decision during the clinical session | | |
| 7 | - | | |

Question 11. Please state to what extent you agree with the following statement: Using the Echo workflow solution can help with an **increased overall echo-lab number of patients** (compared to not having the Echo workflow solution).

| **Response** | | **Number** | **Percent** |
| --- | --- | --- | --- |
| Completely disagree | | 0 | 0% |
| Somewhat disagree | | 0 | 0% |
| Neutral/NA | | 2 | 28% |
| Somewhat agree | | 3 | 43% |
| Completely agree | | 2 | 28% |
| **Comments** | | | |
| 1 | It depends on different factors -> The number of patients do not depend only on faster exam, but also on the hospital and department management of workloads. We cannot say we have 20 more patients. The number of patients may not reflect the workflow faster (see above)  We can say that for a certain type of patients we can do studies with more quality (more 3D echo, more strain) that could not be performed previously because of the lack of time and absence of necessary tools -> some studies could not be done, because additional analysis which take few more minutes could not be done as frequent.  What is the main benefit for these certain patients now that there is a time and tool for doing certain studies?  We believe that we are obtaining more accurate assessments, and therefore decisions based on our reports should be more reliable. | | |
| 2 | Echo is so easy to be done and requested these days that we have a huge waiting list. Because all these new tools and features help everybody to trust Echo results and there is an increased number of patients. | | |
| 3 | It depends on various factors such as the number of sonographers, machines, rooms, etc. | | |
| 4 | To increase the numbers, you must shorten the slot time which people are not very happy to do the same work in less time. Potentially it can improve it because we can accommodate some (urgent) patients at the last minute. | | |
| 5 | With echo workflow solution, you may spend more time for each patient. (Patient registration, ..) but it helps with the quality and patient identification, quality of echo lab. Having Echo workflow solution does not consume so much more time, but you get the benefits from systematic collecting the data and how you are doing reports  If you don’t have the solution, you just bring the patient in and do the echo and patients get out of the lab. | | |
| 6 | Every year we increase the number of studies- it depends on organization. Echo workflow can help with being more efficient. Because of the trust in the tools we can split the work e.g. in the morning simple exams can be done by technicians and complex cases can be done in the afternoon, also we are able to do the reporting in a different time. It helps with workflow plasticity: being flexible and adaptable. | | |
| 7 | - | | |

Question 12. Please state to what extent you agree with the following statement: The Echo workflow solution has led to **improved staff experience by single screen access to the patient data (imaging, and/ or EMR)** (compared to not having The Echo workflow solution)

| **Response** | | **Number** | **Percent** |
| --- | --- | --- | --- |
| Completely disagree | | 0 | 0% |
| Somewhat disagree | | 1 | 14% |
| Neutral/NA | | 0 | 0% |
| Somewhat agree | | 1 | 14% |
| Completely agree | | 5 | 72% |
| **Comments** | | | |
| 1 | You still need to have access to EMR for other information. Large part of data (echo data is in the same screen); it does not prevent opening EMR  But we can focus more on the echo | | |
| 2 | The EMR is not fully integrated. Not all cardiology (cathlab images) are integrated; But still it helps. We also see cardiac CT and MRI images | | |
| 3 | I would like to get the most important data of patient history and know how many images done on the patient. Timeline of different imaging modality would be great and ideally being able to click on patient data and see if patient had an MRI, etc. | | |
| 4 | - | | |
| 5 | Our hospital setting and IT infrastructure would support this, because we put EMR and imaging in the same screen | | |
| 6 | Continuity is felt by Philips solution – good experience is more important for sonographers who are the main users. We have one environment, previously it was different, old fashioned, data on different systems. | | |
| 7 | - | | |

Question 13. Please state to what extent you agree with the following statement: The Echo workflow solution has led to **lower cognitive load for the staff in charge** (compared to not having Echo workflow solution)

| **Response** | | **Number** | **Percent** |
| --- | --- | --- | --- |
| Completely disagree | | 0 | 0% |
| Somewhat disagree | | 0 | 0% |
| Neutral/NA | | 1 | 14% |
| Somewhat agree | | 2 | 29% |
| Completely agree | | 4 | 57% |
| **Comments** | | | |
| 1 |  | | |
| 2 | - | | |
| 3 | - | | |
| 4 | It saves manual tasks. Also for juniors, we can help them easier. Knowing that you can review the complex cases also from the previous exams. | | |
| 5 | It depends on the workflow, when the patient checks in, they are registered and get the ID hospital number; using this number we pull the patient demographic data from hospital system and then staff get to echo lab and they have all the information on screen. This reduces the error. With the help of IT we could do the integration. | | |
| 6 |  | | |
| 7 | - | | |

Question 14. Please state to what extent you agree with the following statement: The Echo workflow solution has led to **better general work experience for the user – user friendliness** (compared to not having Echo workflow solution)

| **Response** | | **Number** | **Percent** |
| --- | --- | --- | --- |
| Completely disagree | | 0 | 0% |
| Somewhat disagree | | 0 | 0% |
| Neutral/NA | | 0 | 0% |
| Somewhat agree | | 2 | 29% |
| Completely agree | | 5 | 71% |
| **Comments** | | | |
| 1 | Example: cardiologists in training (residents) – by moving to these new systems they get in touch and using of this tool very quickly | | |
| 2 | In my opinion its user-friendly- some of my staff think its not user-friendly or complicated at the beginning, but after getting used to it (a couple of weeks) they find it user friendly | | |
| 3 |  | | |
| 4 | User-friendliness helps the experience, what you do every day needs a user friendly and integrated system. | | |
| 5 |  | | |
| 6 | From my point of view everything is similar and user-friendly | | |
| 7 | - | | |

Question 15. Please state to what extent you agree with the following statement: In your opinion and based on your experience, can the Echo workflow solution improve the patient experience **(e.g. less repeated exams, faster diagnosis,..)** (compared to not having The Echo workflow solution)

| **Response** | | **Number** | **Percent** |
| --- | --- | --- | --- |
| Completely disagree | | 1 | 14% |
| Somewhat disagree | | 0 | 0% |
| Neutral/NA | | 0 | 0% |
| Somewhat agree | | 3 | 43% |
| Completely agree | | 3 | 43% |
| **Comments** | | | |
| 1 | There is room for improving this aspect Because many physicians don’t have access to this interface, so we may get extra request for a test that is most probably not necessary (  Additional studies which are not necessary) | | |
| 2 | In my opinion it leads to repeated tests, data transfer is quick | | |
| 3 | There is a general issue (related to [our national] electronic medical record system) because the physician who orders the Echo does not have access to the EMR (immediately or easily) so they don’t know if an Echo was done. They don’t have access to ISCV either (outside of cardiology). During COVID time, we wanted to expose sonographers less to covid patients, an echo was done yesterday but they asked for another echo, and we had to call them to confirm if something new happened that they need a second Echo. | | |
| 4 | Every time there is a request for an echo, we check the previous ones, if the patient had the exam previously, we might just not perform the exam, we repeat less exams. Having that immediate information. It does help us and the patients.  Who can benefit most of this? In-patients who are admitted in the hospital are the ones who need repeated exams in a short period of time. It is very important to have all images and being able to compare them and avoid repeated exams if they are not necessary. | | |
| 5 | Example: a specific type of patient who benefit the most: I can give an example when the patient has Severe pulmonary arterial hypertension and is in ICU and oxygen saturation drops echo has been done what is the cause of desaturation. One possibility is to have PFO (opening between upper chambers). I review the previous exam that shows clearly PFO so there won't be any to repeat the examination so faster diagnosis. | | |
| 6 | In my opinion, maybe they feel better as they spend less time with echo scanning. Probably they have the feeling they have more time with their doctor. They feel confident that we have all the images in our institute. Patients are happy they can have all the information: they always can have a copy of the report in their email | | |
| 7 | - | | |

Question 16. Please state to what extent you agree with the following statement: The Echo workflow solution has led to better **collaboration within your organization (section/department)** (supporting doctors in training/residents in specialization training) (compared to not having Echo workflow solution)

| **Response** | | **Number** | **Percent** |
| --- | --- | --- | --- |
| Completely disagree | | 0 | 0% |
| Somewhat disagree | | 0 | 0% |
| Neutral/NA | | 0 | 0% |
| Somewhat agree | | 2 | 29% |
| Completely agree | | 5 | 71% |
| **Comments** | | | |
| 1 | Residents use this tool for self-training- when they work in echolab get some tools but when they are not working at the echolab they use ISCV for self-training. | | |
| 2 | Within org: Very easy to review images, share the images, from trainees to senior staff,  With other departments: its easy to transfer images. | | |
| 3 | It helps with teaching; they learn how to do measurement and how to report. | | |
| 4 | We collaborate with Intensive Therapy unit (ITU), they perform echo and having a system in common we can share the image. We have better communication with them thanks to the system. They perform and report on the system, and we can see and avoid repeated exams, or They can ask us to look at the exams and for consultation.  Cancer patients come as outpatients. If they did not have done the baseline exam, we do the baseline, otherwise looking at all measurements in the report to see the trend as this information is key as what we are going to say in that report is critical and it might change their treatment. Having the information immediately and it is not just the reporting because of a lot of variation in the operators, you need to see the patient and what we tend to do is to ask someone else to look with you to have 4 eyes instead of two. We are incredibly careful with cancer patients, because you want to assess if there is a change e.g. in GLS | | |
| 5 | Helps the discussion during the multidisciplinary as mentioned earlier | | |
| 6 | In my hospital we work with anesthesia, surgeons, clinicians and internal medicine. In cardiology with distinct roles, we talk the same language since different roles use the same platform the same way. During our meeting we have all the information there, and we just need 10 min. | | |
| 7 | - | | |

Question 17. Please state to what extent you agree with the following statement: The Echo workflow solution benefits **my overall clinical daily work**.

| **Response** | | **Number** | **Percent** |
| --- | --- | --- | --- |
| Completely disagree | | 0 | 0% |
| Somewhat disagree | | 0 | 0% |
| Neutral/NA | | 0 | 0% |
| Somewhat agree | | 0 | 0% |
| Completely agree | | 7 | 100% |
| **Comments** | | | |
| 1 | - | | |
| 2 | Its easier to work, if you work with a solution which is not fast enough or with low quality people get stress during the work. It does benefit the overall | | |
| 3 | - | | |
| 4 | It helps me because of all above mentioned points: user-friendliness, saved reporting time, better communication between units etc.. | | |
| 5 | My daily work/goal is care for patients, it’s about correct diagnosis and appropriate treatment, I will be happy if I can answer the question correctly to deliver the best possible care for the patient. And Echo workflow is helping this task and its relevant for me. Also, this solution is a powerful tool for research.  For instance, when I have one patient coming with shortness of breath and was suspected for mild mitral regurgitation but after reviewing the echo I suspected that it is more sever, we do the TEE and the several mitral regurgitations was clear, which explained the symptoms. | | |
| 6 | Yes- this kind of workflow is good to optimize- we have to optimize to balance quality and quantity | | |
| 7 | - | | |

Question 18. The Echo workflow solution and its benefits (i.e. efficiency improvement and diagnosis accuracy) **might help my department/institution to lower costs** (compared to not having the Echo workflow solution)

| **Response** | | **Number** | **Percent** |
| --- | --- | --- | --- |
| Completely disagree | | 0 | 0% |
| Somewhat disagree | | 0 | 0% |
| Neutral/NA | | 3 | 43% |
| Somewhat agree | | 1 | 14% |
| Completely agree | | 3 | 43% |
| **Comments** | | | |
| 1 |  | | |
| 2 | - | | |
| 3 | I would not call it lowering cost, but it can improve the billing. If you do less unnecessary tests and improve your billing can indirectly reducing costs | | |
| 4 | Time saving is one aspect; however, one important aspect is the outcome of the patients.  A solution/ system that allows you do a better faster diagnosis, it does help patients eventually. That is cost saving as patients are diagnosed quicker and get treated quicker, it helps with long term cost saving.  It makes staff having a better experience and therefore it might help with staff retention | | |
| 5 | The cost must be defined (from what perspective).  From Hospital administration where they have to pay for the installation of echo workflow solution, and pay for maintenance and upgrade, that is a lot, but the cost is justified. It is worth the investment. Having echo workflow solution is necessary and mandatory for echo lab. | | |
| 6 | Time is money but its indirect cost saving. In Private setting, you need to try the cardiology department- organizing the cardiology department efficient work. Have a good machine helps with better and faster diagnosis which may help with having more patients. It does help with the return on investment. | | |
| 7 | - | | |

SECOND ROUND

| **Q6 SECOND ROUND- Which patient group benefits most? And why?** | |
| --- | --- |
| 1 | two types of patients: cardiotoxicity and Heart valvular disease; require more followups-> therefore more consistency needed  further elaboration on benefits: we don’t assess reproducibility between the reader (not implemented in the clinical setting)  Consistency: how similar two readers report  - Structured. Vs traditional report: we all enter the same nomenclature and not everyone use different ways of describing; how we report the mech f vav disease; nc is very heterogenous; the report is focusing on it; standard options for reporting- the same/standard language->  If every clinician use a different terms in the report, you cannot make the decision right away and you may need another exam or imaging modality e.g. MR or CT depends on the disease (full report with all information needed for decision making > more efficient, faster decision making, prevent possible extra exams, if the patient need an intervention, it can start earlier)  -Image acquisition Process is more standardized  Automated analysis Highest reproducibility-> for decision making as you might have sop the chemo or decide. Implementing automated tools helping |
| 2 | - |
| 3 | This applies to every disease for serial echo (it is not for one disease only) and for across the board. |
| 4 | Consistency is also very important: similar way of reporting;  However, a bit of variability is always there but having a template helps for the conditions that require less description, it makes it quicker to report, it becomes more efficient. Of note: free hand typing will remain- you should be also descriptive for complex ones  Imaging and analysis: reproducibility  Imaging analysis with GLS or 3D: reproducibility is higher, operator independent or observer independent, this is however in conjunction with the fact the operator needs to be trained and needs to know where the analysis is not usable because images are poor.  when you have good imaging and good tracking-> more reproducible for serial scans  Patients requiring serial scans e.g. cancer patients, as any change in values can have an impact on the treatment  having a good software recognizing and tracking well, compare serial scans-> reproducible |
| 5 | Congenital heart disease, everything is mentioned in the report in a consistent way. We have referred patient we send the report back. We customized the report in my lab |
| 6 | Reproducibility and consistency are important for chronic patients who require serial measurements and comparison of exams needed, as you need to repeat the same type of study to compare the same parameters overtime.  For example, every time you check EF for patients with heart failure  Or for Patients with atrial fibrillation, you always measure the left atrium diameter.  Cardio oncology patients  Patients with heart failure  Patients with assistance devices  Pulmonary hypertension (1-2 echos every year)  Ranking: especially for patients that EF is critical. Patients with heart failure or with transplantation  Benefit of using the same language in reports in communication and collaboration (e.g. with referrals)  Its key to use more or less common language in reporting, patients with heart failure, we try to combine all information needed for 1)clinical aspects (to make decision, e.g. we describe mitral valve disease always the same way in Echo lab which is important for intervention Cath lab and surgeon, because they understand all about the report in mitral valve), 2)teaching, 3)research |
| 7 | Patient with complex cardiac diseases or abnormal cardiac anatomy |

| **Q9 SECOND ROUND Which patient group benefits most? And why?** | |
| --- | --- |
| 1 | Patients who need repeated echos, history and overview of patients; we are more accurate in scheduling the patients  Same echo number -> more time for postprocessing  Has decision making changed based on the strain? We don’t have an answer to that- To be Measured by echocardiologist and Oncologist or cardioncologist |
| 2 | - |
| 3 | This applies to every disease for serial echo (it is not for one disease only) and for across the board. I look for every patient who are going through Echo Imaging. |
| 4 | Again for cancer patients: immediate detection of change; reduction of GLS  Massive vegetation of heart valve, if you have a very good 3D on transthoracic (TTE): you could potentially save the patient from further imaging TOE/TEE; If you send the patient for TOE/TEE-> it delays the diagnosis. Via a good 3D on TTE on mitral valve you clearly see two massive vegetation so TTE was good enough 3D on TTE: has limitations: on normal structure or poor picture I would never do it, but if I have a good image, suspicion of mass vegetation, prosthetic valve, but not many do it on TTE |
| 5 | When my team does the echo and are not sure and they need more consultation, this solution allows the consultation from anywhere  Accuracy is higher and time to diagnosis  Sometimes an interventionist needs more consultation (between departments) if they are not certain.  We have complex patients with multiple problems and morbidities that need consultation and this solution help.  Patients with covid 19, usually the question was if it is pneumonia or pulmonary edema. After doing the echo t depends a lot on workflow, they were not sure if the patient has a normal function, critical care physician could us ethe report and look at the eco to manage patient correctly. If you don’t have this workflow the consultation was very difficult.  Flexibility: the solution helps with being flexible for scheduling the patient based on their complexity |
| 6 | 1-Time is critical for fast diagnosis for Intensive care patients  2-Complex patients: in valvular heart disease when there is more than 1 valve involved at the same time. Patients with pulmonary hypertension as you need to focus on the right ventricle, sometime right ventricle is very complex and we need to use all the tools in our system  3-cancer patients are not complex as Ejection fraction is needed. Sometimes you have the echo in the morning and on the same day the patient goes to the oncologist, so the report should be ready very fast.  In my hospital the majority (90%) are complex cases (heart failure, with valvular disease, pulmonary hypertension). In my clinic 75% are complex. |
| 7 | - |

| **Q10 SECOND ROUND Which patient group benefits most? And why?** | |
| --- | --- |
| 1 | Decision making has been enhanced in any scenario that needs several measurements, and/ or require consistency and comparison with previous measurements. Examples are heart valvular disease, adult congenital disease and cancer patients.  The more complex the scenario the greater the benefit of this solution. In case of heart valvular disease you need 20-40 measurements, another indication is adult congenital disease and our center is the national reference center for this kind of disease,  The solution allows to be more consistent  20% of echos in our center are adult congenital disease, we do a lot of echos on these patients (referred by hospitals that are 100-200km away) previously we could not capture how to report on such complex patients, a lot of information measurements,  Most of time these reports are interpreted in our center-> now we have a common language to report; these can be taken to other hospitals, the key information should be in these reports-> Crystal clear report on condition and status of patient  Efficient communication between doctors? Yes, Between my center and referring physicians (receiving hospitals) taking back the patient  The system itself is the reference (common place) for the information- in the past all echo info was embedded scattered in electronic medical records  Cardiotoxicity more homogenous; Heart valvular disease more heterogenous |
| 2 | - |
| 3 | This applies to every disease for serial echo (it is not for one disease only) and for across the board. Rapid access to information is necessary for all patients.  Another question is if junior sonographer/echocardiologist can benefit more than senior ones?  This will be beneficial for both junior and senior sonographers/echo cardiologist. |
| 4 | Sometimes you have an idea about the patient based on the clinical image, you do the study: based on the patient presentation and clinical scenario you are suspicious of something you go to your imaging and do advanced imaging as well that fits with the clinical picture (your initial idea), that makes you more confident. Without doing extra imaging.  It helps with flexibility- You can do postprocessing during study if you got time, otherwise you can do it later. Also if junior wants to do postprocessing with some one senior its more flexible  Familiarity with the advanced imaging tech, how easy is to take the image: I have to make a quick decision and need to spend 15 min on the imaging- I won't do it to be quick.  While I have to press a button so it’s easy, then people may realize that despite the time pressure, but they get information very quickly, then they will do it maybe. Philips new TT software is like having a button.  in terms of being confident you need something user-friendly, that is reliable and gives you the right information quickly. That gives you immediate idea of what is happening- with this it would definitely increase your confidence as it gives you additional information doing the postprocessing and it adds up to your study. GLS software of Philips 7-8 years ago it took 20 min for postprocessing and you don’t have 20 min  you can do the Postprocessing another time: flexibility  Junior can do the postprocessing with senior; I disagree with having division based on experience, only at beginning of echo training is ok. |
| 5 | You have the record of patients and you have the modalities and tools and findings, you send the patient for surgery or intervention and they find out something that it was in addition to what you diagnosed, so it is strong point for learning, helping with feedback loop, If the patient has additional information e.g. CT scan, surgery you can look back at echo again and identify what has been missed initially or what should be cautious about. You can build databases with Reference patients and examples which can be beneficial for next generation and used for teaching.  For Complex cases it can be beneficial. A patient who has sever stenosis and was referred because of his chest pain. He was sent for echo the echo showed that aortic valve stenosis is not sever, the patient was sent the patient back because ethe Dr thought the stenosis is not sever but need follow up on patient, when we read the echo, although the patient had coronary artery disease and coronary angiogram was needed, review aspect of this solution help with such a situation. |
| 6 | Again, its about complex cases as these are the ones that need all your tools. You need to feel confident with complex cases, to be confident for these cases you need to be a good cardiologist but also you need to have good tools. Asking for others opinion: for complex cases  Communication is important for complex cases. I receive requests from others to review the cases with them. Its of note that even a senior cardiologist with years of experience need good tools to feel confident about their decisions.  Juniors need training and learning curve is Improving because of tools. As a fellow you have all the information available, and you can use the available tools and information and you can train yourself |
| 7 | - |
